# Clinical and Epidemiological Characteristics of the First Month of the Covid-19 Pandemic in Chile

**DOI:** 10.1101/2020.08.11.20171975

**Authors:** Macarena R. Vial, Anne Peters, Inia Pérez, María Spencer, Mario Barbé, Lorena Porte, Thomas Weitzel, Mabel Aylwin, Pablo Vial, Rafael Araos, Jose M. Munita, for the CAS-UDD Covid-19 Working Group

**Author notes:** Corresponding author: Jose M. Munita. Alternate corresponding author: Rafael Araos. Clínica Alemana - Universidad del Desarrollo Covid-19 working group: Alejandra Marcotti, Jorge Pérez, Luis Miguel Noriega, Pablo Gaete, Sebastián Solar, Silvina López, Paulette Legarraga, Valeska Vollrath, Alicia Anderson, Mirentxu Iruretagoyena, Jerónimo Graf, Rodrigo Pérez, Manuela A. Roa.

## Abstract

**Introduction:** Understanding the clinical course and outcomes of patients with Covid-19 in underrepresented populations like Latin America is paramount. In this study, we report the clinical characteristics of Covid-19 in Chile, with a focus on subjects requiring hospitalization during the initial phases of the SARS-CoV-2 pandemic.

**Methods:** This is a single center study including all consecutive patients diagnosed with Covid-19 during the first month of the pandemic. Demographics, clinical characteristics and laboratory data were collected within 24 hours of admission. The primary outcome was a composite of ICU admission or all-cause, in-hospital mortality.

**Results:** During the first month of the pandemic, 381 patients were confirmed as positive for SARS-CoV-2 by molecular testing; 88 (23.1%) of them eventually required hospitalization. Median age of the cohort was 39 years (IQR 31-49). Overall mortality was 0.7% and 18 (3.7%) out of the 88 subjects who required hospitalization either died and/or required ICU. Increased body mass index (BMI), C-reactive protein levels (CRP) and the SaTO2/FiO2 index on admission were independently associated with a higher risk of ICU care or death.

**Discussion:** The lower mortality observed in our prospective cohort during the first month of SARS-Cov-2 pandemic was lower than previously reported. This finding could be due to a lower threshold for admission, a healthcare system not yet overburdened and a younger population, among other factors. BMI, CRP on admission were strong predictors for ICU care or all-cause, in-hospital mortality. Our data provide important information regarding the clinical course of Covid-19 in Latin America.

## Introduction

In December 2019, a cluster of severe pneumonia of unknown etiology was reported in Wuhan, Province of Hubei, China.^1^ A rapid investigation determined that the agent involved was a novel Coronavirus sharing significant sequence identity with bats and human-related coronaviruses.^2,3^ The virus and its associated disease were named <u>S</u>evere <u>A</u>cute <u>R</u>espiratory <u>S</u>yndrome <u>Co</u>rona<u>v</u>irus <u>2</u> (SARS-CoV-2) and <u>C</u>orona<u>v</u>irus <u>d</u>isease 19 (Covid-19), respectively. ^4^ Soon after its identification, SARS-CoV-2 spread globally causing tremendous burden to health care systems and society as a whole. According to publicly available information, the first case of Covid-19 in Chile was diagnosed on March 3, 2020. Initial cases were imported from Europe, but family clusters and evidence of secondary transmission were rapidly observed. Therefore, the Chilean government declared phase 4 (widespread, ongoing local transmission) on March 16, 2020. Data from early clinical series have shed light on the clinical presentation of Covid-19. After a median incubation time of 5 days, ^5,6^ patients developing symptoms usually present with an influenza-like illness, with fever, dry cough, headache, odynophagia, and dyspnea as the most common symptoms^1^. Interestingly, a large proportion of subjects also report the development of anosmia and/or augesia as one of the cardinal symptoms of the infection. Early work from China showed that 80% of the subjects with Covid-19 followed an uncomplicated course, 15% required hospital admission and 5% developed a severe infection with a catastrophic respiratory failure needing critical care support. ^5^ The case-fatality rate is variable, ranging from 2.25% in the Republic of Korea to 14.5 % in countries like Italy, with increasing age, and comorbidities being the most important predictors of mortality.^7,8^ As Covid-19 spreads, and the number of cases dramatically increases, a detailed description of Covid-19 in new geographical areas is critical. Moreover, understanding the clinical course and outcomes of patients with Covid-19 in underrepresented populations like Latin America is paramount. In this study, we report the clinical characteristics of Covid-19 in Chile with a focus on subjects hospitalized during the first month of the epidemic.

## Methods

The study was conducted in Clínica Alemana de Santiago (CAS), a tertiary care not-for-profit hospital in Santiago, Chile. CAS is a 442-beds healthcare facility that before the Covid-19 pandemic included 12 general and 10 cardiac intensive care unit (ICU) beds. The study was approved by the Institutional Review Board, with a waiver of informed consent given that data was collected as part of routine clinical practice and used in a de-identified manner. All consecutive patients attending the Emergency Room and diagnosed with Covid-19 between March 3, 2020, and April 4, 2020, were included in this cohort. Laboratory testing for SARS-CoV-2 infection was performed using a commercial reverse-transcriptase (RT) polymerase chain reaction (PCR) of nasopharyngeal and oropharyngeal swab samples.^9^ Cases were identified using the central laboratory database and electronic medical records. Only patients with a positive SARS-CoV-2 RT-PCR were included. Data was extracted from the electronic medical records and entered into a REDCap database by a team of researchers, after a training session led by the senior data manager investigator (AP). Two study investigators (MS and AP) conducted weekly data audits to ensure the quality of the collected data. Data collected included demographic information, comorbidities, clinical presentation, as well as duration of symptoms, treatment, and outcomes. Laboratory values were automatically extracted from the electronic medical records. The Charlson Comorbidity Index (CCI) was used to summarize comorbidities. The score goes from 0 to 24, with zero representing no comorbidities.^10^ Body Mass Index (BMI) was used to assess excess body weight. Pulse oximetry saturation (SpO_2_)/ Fraction of inspired oxygen (FiO_2_) ratio was used to assess respiratory exchange.

The primary outcome was a composite of ICU admission or all-cause, in-hospital mortality. Variables used to assess risk factors for the main outcome were obtained within 24 hours of admission.

### Statistical analysis

Continuous and categorical variables were presented as median (IQR) and n (%), respectively. Patient characteristics were compared by subgroups of interest using a chi-square test, Fisher’s exact test or the Mann-Whitney U test as appropriate. A two-tailed p-value <0.05 was considered statistically significant for all analyses.

To explore risk factors associated with disease severity, univariable and multivariable logistic regression models were used. We selected variables for the logistic regression model based on previous findings and plausibility. A maximum of 8 variables were included in the logistic regression model to avoid overfitting. All analyses were conducted using Stata (Version 16.0. College Station, TX: StataCorp LLC).

## Results

During the first month of the Covid-19 pandemic, 3679 subjects presented with possible SARS-CoV-2 infection and tested by RT-PCR in our institution; 381 (10.4%) were confirmed as positive and included for this study. The average time of symptoms at the time of initial testing was 3.7 days (SD 3.8).

The median age of the cohort was 39 years (IQR 31-49) and 153 (52%) were female (Table 1). A total of 253 (66.4%) patients reported an epidemiological risk factor for acquiring SARS-CoV-2, i.e. 206 had exposure to a confirmed case, and 47 had recently visited a highrisk country, (16 reported both risk factors). Cough and fatigue were the most common symptoms at presentation. A summary of the main demographic and clinical features of the cohort is shown in Table 1.

**Table 1.** Demographic and clinical presentation of patients with Covid-19 diagnosed during the first month of the SARS-CoV-2 pandemic.

|  | **All outpatients**<br>**N=293** | **All inpatients**<br>**N=88** | **p-value** |
| --- | --- | --- | --- |
| **Characteristic** |  |  |  |
| Age, years | 37 (28-45) | 49 (39.5-65) | .00 |
| Female gender | 153 (52.2) | 45 (51.1) | .85 |
| **Comorbidities** |  |  |  |
| Diabetes | 5 (1.7) | 6 (6.8) | .73 |
| Hypertension | 15 (5.1) | 25 (28.4) | .00 |
| BMI ≥ 30 | 3 (1) | 14 (15.9) | .002 |
| Current smoker | 7 (2.4) | 6 (6.8) | .055 |
| Obstructive pulmonary disease | 10 (3.4) | 6 (6.8) | .17 |
| Malignancy | 3 (1) | 3 (3.4) | .14 |
| Mean CCS (SD) | 0.28 (0.7) | 1.38 (2.0) | .00 |
| **Symptoms at the time of initial presentation** |  |  |  |
| Cough | 170 (58) | 64 (72.7) | .013 |
| Fever | 167 (43.8) | 69 (78.4) | .00 |
| Odynophagia | 126 (43) | 35 (39.8) | .59 |
| Fatigue | 91 (31.1) | 38 (43.2) | .036 |
| Anosmia or Ageusia | 28 (9.6) | 7 (8) | .65 |
| Chest pain | 9 (2.4) | 5 (5.7) | .26 |
| Dyspnea | 3 (1.0) | 4 (4.5) | .048 |

Data are median (IQR), n (%), or n/N (%). p values were calculated by Mann-Whitney U test, $\chi^2$ test, or Fisher's exact test, as appropriate. $\chi^2$ test comparing all subcategories.

**Table 2.** Baseline characteristics at admission of patients who required ICU care or died and those who did not.

|  | All hospitalized N=88 | ICU care or death N=18 | Non-ICU care N=70 | p-value |
| --- | --- | --- | --- | --- |
| **Demographics** |  |  |  |  |
| Age, years | 49 (39.5-65) | 68.5 (59-72) | 46 (38-58) | .000 |
| Female gender | 45 (51.1) | 3 (16.7) | 42 (60) | .003 |
| **Symptoms on admission** |  |  |  |  |
| Cough | 64 (72.7) | 13 (72.2) | 51 (72.9) | .96 |
| Fever | 69 (78.4) | 15 (83.3) | 54 (77.1) | .64 |
| Anosmia | 7 (8) | 0 | 7 (10) | .16 |
| Ageusia | 3 (3.4) | 0 | 3 (4.3) | .37 |
| Odynophagia | 35 (39.8) | 7 (38.9) | 28 (40) | .93 |
| Fatigue | 38 (43.2) | 6 (33.3) | 32 (45.7) | .35 |
| Dyspnea | 4 (4.5) | 2 (11.1) | 2 (2.9) | .16 |
| Chest pain | 5 (5.7) | 0 | 5 (7.1) | .24 |
| **Comorbidities** |  |  |  |  |
| Diabetes | 6 (6.8) | 3 (16.7) | 3 (4.3) | .06 |
| Hypertension | 25 (28.4) | 10 (55.6) | 15 (21.4) | .004 |
| Mean BMI (SD) | 26.5 (3.8) | 29.2 (3.9) | 25.7 (3.5) | .002 |
| Smoker | 6 (6.8) | 1 (5.6) | 5 (7.1) | .81 |
| COPD or Asthma | 6 (6.8) | 3 (16.7) | 3 (4.3) | 0.097 |
| Mean CCS (SD) | 1.38(1.96) | 3(2.3) | 0.96 (1.62) | .001 |
| **Physical exam** |  |  |  |  |
| Heart rate | 81 (74-89) | 86 (75-97) | 80.5(74-88) | .23 |
| T >37.5 C (axillary) | 42/48 (87.5) | 8/9 (88.9) | 34/39 (87.2) | 0.89 |
| Respiratory rate | 20 (18-22) | 21(20-24) | 20(18-22) | .13 |
| SBP <90 Or MAP < 65 mmHg | 9/81 (11.1) | 1/13 (7.7) | 8/68 (11.8) | 1.0 |
| SaTO2%/FiO2 | 452.4 (404.2-461.9) | 317.9 (248.6-447.6) | 457.14 (447.6-466.7) | .000 |
| **Chronic Medications** |  |  |  |  |
| ARBs/ACEi | 18/87 (20.7) | 9/17 (52.9) | 9/70 (12.9) | .00 |
| Steroids | 4 (4.5) | 4 (22.2) | 0 | .001 |

### Subgroup of patients requiring hospitalization

Among the 381 patients, 88 (23.1%) eventually required hospital admission, 51 of them were hospitalized after an initial management as outpatients. The median time of symptoms at the time of admission was 8 days (IQR 5-10). Hospitalized patients were generally older than those managed as outpatients (median 49 vs. 37, p < 0.001) and had a higher CCS with 0.28 for outpatients and 1.38 for inpatients (p < 0.001). On the first day of hospitalization, 40 (45%) subjects were admitted to general wards, 41 (47%) to a step-down unit, and 7 (8%) to ICU. Among subjects hospitalized in general wards and stepdown units, 10 (12%) patients were subsequently transferred to the ICU. The median duration of hospitalization was 8 (4-14.5) days and the median length of stay (LoS) in the ICU was 13 (5.3-17.8) days. A total of 82 (93%) patients had an imaging compatible with pneumonia; 39 (44%) required supplementary O_2_ only, 20 (23%) non-invasive ventilation and 10 (11%) invasive ventilation. Median days of invasive ventilation were 7.5 (6.3-15.3). Among those hospitalized, excess body weight (62.5%) and hypertension (28%) were the most common coexisting conditions A summary of the laboratory abnormalities on admission is presented in Table 3 and Figure 1. The most common laboratory abnormalities were: lymphopenia, increased levels of C-reactive protein (CRP) and D-dimer. Ferritin values were available for 25 patients at admission, with 20 (80%) presenting an increased level. A comparison of patients who presented severe disease, defined as need for ICU care at any time during admission or inhospital death, with non-severe disease is provided in Table 3. In univariate analysis, odds of ICU care/death were higher in males, and those with higher BMI, older age, a history of diabetes or hypertension, and chronic medication such as steroids, angiotensin-converting enzyme (ACE) inhibitors or angiotensin II receptor blockers (ARBs). Increased white blood cells, neutrophils, CRP, procalcitonin, ferritin, D-dimer, bilirubin, troponin T, LDH and lower SaTO2/FiO_2_, prothrombin time, sodium, albumin and lymphocytes were all associated with the need for ICU care or death. Based on clinical plausibility and significance level on the univariate analysis, the following variables were included in the multivariate logistic regression model: age, gender, CRP, BMI, neutrophil lymphocyte ratio, SaTO2/FiO_2_, D-dimer and sodium. Increased BMI and CRP levels were independently associated with increased odds of ICU care/death. SaTO_2_/FiO_2_ had a p-value of 0.051, not significant according to our predetermined level of 0.05. See Table 3. As of July 31, out of the 88 patients who required hospitalization, a total of 3 patients had died and 85 had been discharged and remained alive.

**Figure 1:**
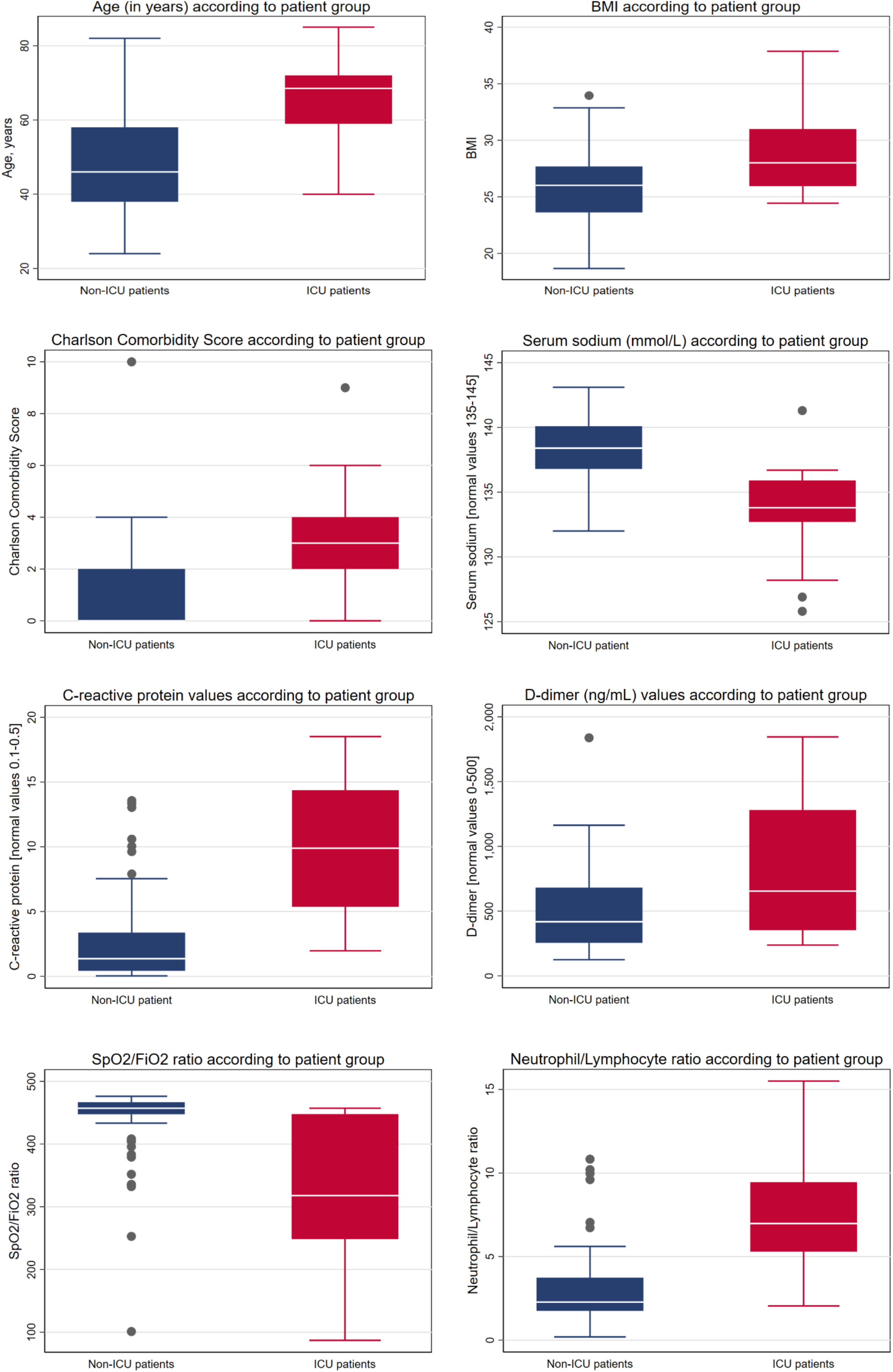
Clinical and laboratory markers within 24 hours of admission comparing ICU vs non-ICU patients.

**Table 3:** Laboratory findings within 24h of admission comparing patients who required ICU care or died to those who did not require ICU

|  | All patients | ICU care or death | No ICU care | p-value |
| --- | --- | --- | --- | --- |
| Hemoglobin, g/dL | 14.1 (13.1-14.8) | 10.2(10.1-12.5) | 14.3 (13.5-15) | .13 |
| White blood cell count, mm<sup>3</sup><br>< 4500<br>> 11500 | 23/73 (31.5)<br>2/73 (2.7) | 1/15 (6.7)<br>1/15 (6.7) | 22/58 (37.9)<br>1/58 (1.7) | .096<br>.024 |
| Lymphocyte count, mm<sup>3</sup><br>< 1000 | 31/73 (42.5) | 12/15 (80) | 19/58 (32.8) | .001 |
| Neutrophil count, mm<sup>3</sup><br><2500 | 3605 (2453-4714)<br>19/72 (26.4) | 4723 (4095-6707)<br>0 | 3266 (2304-4192)<br>19/57 (33.3) | .006<br>.013 |
| Neutrophil/Lymphocyte ratio | 2.8 (1.9-5.5) | 7.0 (5.3-9.4) | 2.3 (1.8-3.7) | .00 |
| Platelet count, mm<sup>3</sup><br>< 100 | 182(136-236)<br>2/73 (2.7) | 175(131-252)<br>0 | 186 (136-236)<br>2/58 (3.4) | .58<br>.067 |
| C-reactive protein mg/dL >0.5 | 5/73 (75.3) | 15/15 (100) | 40/58 (69) | .015 |
| Procalcitonin ng/mL | 0.05 (0.04-0.1) | 0.17 (0.08-0.3) | 0.05 (0.03-0.07) | .003 |
| Ferritina ng/mL > 300 | 20/25 (80) | 11/11 (100) | 9/14 (64.3) | .046 |
| D-dimer, ng/mL > 500 | 32/68 (47.1) | 10/14 (71.4) | 22/54 (40.7) | .040 |
| Prothrombin time, s | 88.5 (81-99.5) | 85 (73-87) | 92 (85-100) | .014 |
| Aspartate aminotransferase, U/L | 27.3 (21.1-35) | 26.5 (23.2-42) | 27.3 (20.9-35) | .90 |
| Alanine aminotransferase, U/L | 26.1(16.4-38.6) | 28.3 (15.2-38.6) | 25.7 (16.4-40) | .54 |
| Total bilirubin, mg/dL | 0.36 (0.25-0.48) | 0.51 (0.38-0.7) | 0.33 (0.24-0.42) | .001 |
| Albumin, g/dL | 4.05 (3.8-4.4) | 3.7 (3.4-4.1) | 4.1 (3.9-4.4) | .003 |
| Lactate dehydrogenase U/L | 222 (177-296) | 264 (203-411) | 205 (171-275) | .011 |
| Troponin T, ng/mL<br>> 14 | 3.2 (3-7.3)<br>7/46 (15.2) | 13.3 (4-30.2)<br>5/11 (45.5) | 3 (3-5.3)<br>2/35 (5.7) | .043<br>.005 |
| Sodium mmol/L < 135 | 18/77 (23.4) | 11/18 (61.1) | 7/59 (11.9) | .00 |
| Creatinine, mg/dL | 0.80 (0.72-0.97) | 0.87 (0.73-0.99) | 0.78 (0.72-0.93) | .93 |

Data are median (IQR) or n/N where N is the total number of patients with available data.

**Table 4.** Multivariate Logistic Regression Analysis Risk Factors for ICU or death for patients admitted with COVID-19

|  | OR (95% CI) | p-value |
| --- | --- | --- |
| BMI | 1.436 (1.042-1.979) | 0.027 |
| C-reactive protein | 1.505 (1.107-2.047) | 0.009 |

## Discussion

Chile has been one of the most affected countries by the spread of SARS-Cov-2 worldwide, and still struggling to contain the first wave of the Covid-19 pandemic. We report the first prospective cohort of patients with Covid-19 in Chile and one of the few available for Latin America and other developing regions. The 381 patients of our cohort encompass all patients diagnosed at our institution during the first month of the pandemic and represent a significant proportion of all notified cases in Chile during the study period (n = 4,161). The cohort mirrored the beginning of the epidemic curve (first four weeks), when a large proportion (~65%) of patients remained able to identify a risk of exposure to SARS-CoV-2; thus, many subjects attended the hospital despite having minor symptoms to receive advice and obtain a diagnosis. Importantly, in Chile, summer vacations go from December to the first week of March, so despite frontiers closing on March 16 (only 13 days after the first case), many cases were brought into the country by returning travelers. Indeed, 47 (12.3%) of patients in our cohort had recently visited what was considered at the time a high-risk country. These data suggests that strict isolation and contact tracing of imported cases may have resulted in decreased spread to the general population.

A large proportion of patients in our cohort had a mild presentation, with only 18 (4.7%) out of 381 requiring ICU care, consistent with prior studies.^11^ Interestingly, only 3 patients died during the hospitalization, representing a 3.4% of all patients requiring admission and 0.7% of the overall cohort. An in-hospital mortality rate of 3.4% is strikingly lower compared to previous large reports from Wuhan (28%), the New York Area (21%) and other European countries.^12–15^ This difference could be explained by a lower threshold for admission in our cohort at a time when the healthcare system was not yet overloaded and bed capacity was high. However, an in-hospital mortality of 22% reported from Germany at a time when the healthcare capacity was not burdened argues against this as the sole explanation.^14^ Also, patients in our cohort were younger (median age 49) compared to the New York, Wuhan and German cohorts (median ages 63, 56 and 72 respectively). Age has been consistently associated with disease severity and outcomes ^12,16^, hence, it is likely a contributing factor in the lower mortality rates observed in our cohort. Finally, previous cohorts ^14,15^ excluded a large proportion of patients who were still hospitalized at the time of study closure, biasing results towards an increased mortality rate due to the higher inclusion of patients who died early in the course of hospitalization

The cohort presented here is mostly Hispanic, an ethnic population generally underrepresented in medical research.^17^ Data from the United Kingdom and USA has shown an increased risk of severe COVID-19 among ethnic minorities.^18–20^ The low fatality and ICU rate of our cohort suggests that the ethnicity issue might rather be a disparity problem. Studies appropriately representing minority populations are sorely needed. Indeed, failing to include an ethnically representative population leads to results that may not apply to these groups, increasing the health inequality.

As reported elsewhere,^21^ higher CRP levels were associated with increased odds of need for ICU care or in-hospital death. Further, our data suggest that elevated CRP levels in the first 24 hours of admission were a biomarker for severe clinical presentation of Covid-19. CRP is an acute phase protein released mainly in response to interleukin-6 (IL-6), a cytokine that has also been associated with disease severity in Covid-19.^12^ Higher CRP levels are likely associated with a higher inflammatory response, which may correspond with increased tissue damage. CRP levels are widely available and generally cheaper than measuring IL-6 levels and therefore may represent an interesting biomarker to investigate in future studies. Increased BMI was also associated with severe disease. Several theories have been raised to explain this association, which was described before in other cohorts^22^, including overactivated inflammation and immune response, decreased chest expansion, and increased expression of ACE 2, among others.^22–23^ Although the mechanisms are beyond the scope of this study, BMI seems to be a useful predictor of Covid-19 severity and therefore obese patients should be considered a high-risk population. Finally, although the SaTO_2_/FiO_2_ index on admission was not independently associated with our main outcome (p=0.051), it was very close to our pre-established level of significance. These data, along with a high biological plausibility suggests this index is worth exploring as a marker for the development of severe Covid-19 disease in future studies.

The role of other biomarkers previously identified as predictors of in-hospital mortality or ICU need were not confirmed in our cohort. These findings could be explained due to the limited number of patients included in our series along with the low frequency of occurrence of our primary outcomes (i.e. ICU admission and/or in-hospital mortality). In addition, other relevant limitations of our study include that it is a single center effort and its observational nature. Due to the latter, we did not explore treatment effects given our limited ability to appropriately correct for potential confounders. However, our data were prospectively collected with high quality standards and provide one of the few studies contributing information from developing areas of the world, in this case South America. As mentioned above, the data of this cohort mainly represent the initially affected high-income population of Chile and a time of the pandemic were the healthcare system was not yet overwhelmed. Therefore, future studies analyzing the general population attending to a wider range of hospital centers and reflecting a systemic stress created by the large number of patients infected with SARS-CoV-2 will be important to help understand the possible influence of social and health disparities, and of the system overload in the outcomes of Covid-19 patients.

In conclusion, among patients in our study, SARS-CoV-2 generally caused mild illness with a case fatality rate of 0.7%. On admission, variables associated with the need of ICU care and/or in-hospital mortality included BMI, CRP and SaTO2/FiO_2_, all of which are widely available in low and middle resource settings such as Latin America.

## Data Availability

Please contact the corresponding author

## Author contribution

MRV: data analysis and interpretation, writing the original draft; AP: acquisition of data and data analysis; IP: study design and critical review of the manuscript; MS: study design and data acquisition; MB: study design and data acquisition; LP: data acquisition and critical review of the manuscript; TW: study design and critical review of the manuscript; MA: study design, data acquisition; PV: study design and critical review of the manuscript. RA: study design, writing the original draft and critical review of the manuscript; JMM: conception and design of the study design, critical review of the manuscript and final approval of the version to be submitted.

## Acknowledgments

We would like to thank Betel Rivero, Patricia Vargas, Marco Maldonado and Magdalena Canals for their valuable help with data collection and the Departamento Cientifico Docente from Clinica Alemana de Santiago for their constant support.

## References

1- Huang C, Wang Y, Li X, et al. Clinical features of patients infected with 2019 novel coronavirus in Wuhan, China [published correction appears in Lancet. 2020 Jan 30;]. Lancet. 2020; 395(10223):497–506

2- Zhou P, Yang XL, Wang XG, et al. A pneumonia outbreak associated with a new coronavirus of probable bat origin. Nature. 2020;579(7798):270–273.

3- Zhu N, Zhang D, Wang W, et al. A Novel Coronavirus from Patients with Pneumonia in China, 2019. N Engl J Med. 2020; 382(8):727–733

4- Coronaviridae Study Group of the International Committee on Taxonomy of Viruses. The species Severe acute respiratory syndrome-related coronavirus: classifying 2019-nCoV and naming it SARS-CoV-2. Nat Microbiol. 2020; 5(4):536–544

5- Guan WJ, Ni ZY, Hu Y, et al. Clinical Characteristics of Coronavirus Disease 2019 in China. N Engl J Med. 2020; 382(18):1708–1720

6- Lauer SA, Grantz KH, Bi Q, et al. The Incubation Period of Coronavirus Disease 2019 (COVID-19) From Publicly Reported Confirmed Cases: Estimation and Application. Ann Intern Med. 2020; 172(9):577–582

7- Du RH, Liang LR, Yang CQ, et al. Predictors of mortality for patients with COVID-19 pneumonia caused by SARS-CoV-2: a prospective cohort study. Eur Respir J. 2020;55(5):2000524. Published 2020 May 7.

8- Wang K, Zuo P, Liu Y, et al. Clinical and laboratory predictors of in-hospital mortality in patients with COVID-19: a cohort study in Wuhan, China [published online ahead of print, 2020 May 3]. Clin Infect Dis. 2020; ciaa538.

9- Porte L, Legarraga P, Vollrath V, et al. Evaluation of novel antigen-based rapid detection test for the diagnosis of SARS-CoV-2 in respiratory samples [published online ahead of print, 2020 Jun 1]. Int J Infect Dis. 2020;S1201-9712(20)30405–7

10- Charlson ME, Pompei P, Ales KL, MacKenzie CR. A new method of classifying prognostic comorbidity in longitudinal studies: development and validation. J Chronic Dis. 1987;40(5):373–383.

11- Wang D, Hu B, Hu C, et al. Clinical Characteristics of 138 Hospitalized Patients With 2019 Novel Coronavirus-Infected Pneumonia in Wuhan, China [published online ahead of print, 2020 Feb 7]. JAMA. 2020;323(11):1061–1069

12- Zhou F, Yu T, Du R, et al. Clinical course and risk factors for mortality of adult inpatients with COVID-19 in Wuhan, China: a retrospective cohort study [published correction appears in Lancet. 2020 Mar 28;395(10229):1038] [published correction appears in Lancet. 2020 Mar 28;395(10229):1038]. Lancet. 2020;395(10229):1054–1062.

13- Richardson S, Hirsch JS, Narasimhan M, et al. Presenting Characteristics, Comorbidities, and Outcomes Among 5700 Patients Hospitalized With COVID-19 in the New York City Area [published online ahead of print, 2020 Apr 22] [published correction appears in doi: 10.1001/jama.2020.7681]. JAMA. 2020;323(20):2052–2059.

14- Karagiannidis C, Mostert C, Hentschker C, et al. Case characteristics, resource use, and outcomes of 10 021 patients with COVID-19 admitted to 920 German hospitals: an observational study [published online ahead of print, 2020 Jul 28]. Lancet Respir Med. 2020;S2213-2600(20)30316–7.

15- Docherty AB, Harrison EM, Green CA, et al. Features of 20 133 UK patients in hospital with covid-19 using the ISARIC WHO Clinical Characterisation Protocol: prospective observational cohort study. BMJ. 2020;369:m1985

16- Du RH, Liang LR, Yang CQ, et al. Predictors of mortality for patients with COVID-19 pneumonia caused by SARS-CoV-2: a prospective cohort study. Eur Respir J. 2020; 55(5):2000524.

17- Smart A, Harrison E. The under-representation of minority ethnic groups in UK medical research. Ethn Health. 2017;22(1):65–82Smart A, Harrison E.

18- Khunti K, Singh AK, Pareek M, Hanif W. Is ethnicity linked to incidence or outcomes of covid-19?. BMJ. 2020;369:m1548. Published 2020 Apr 20

19- Public Health England. Disparities in the risk and outcomes of COVID-19. 2020. https://assets.publishing.service.gov.uk/government/uploads/system/uploads/attachment_data/file/890258/disparitiesreview.pdf

20- Haynes N, Cooper LA, Albert MA; Association of Black Cardiologists. At the Heart of the Matter: Unmasking and Addressing the Toll of COVID-19 on Diverse Populations. Circulation. 2020;142(2):105–107.

21- Luo X, Zhou W, Yan X, et al. Prognostic value of C-reactive protein in patients with COVID-19 [published online ahead of print, 2020 May 23]. Clin Infect Dis. 2020.

22- Yang J, Hu J, Zhu C. Obesity aggravates COVID-19: a systematic review and meta-analysis [published online ahead of print, 2020 Jun 30]. J Med Virol. 2020;10.

23- Simonnet A, Chetboun M, Poissy J, et al. High Prevalence of Obesity in Severe Acute Respiratory Syndrome Coronavirus-2 (SARS-CoV-2) Requiring Invasive Mechanical Ventilation. Obesity (Silver Spring). 2020;28(7):1195–1199.

